# Comparing Metapopulation Dynamics of Infectious Diseases under Different Models of Human Movement

**DOI:** 10.1101/2020.04.05.20054304

**Authors:** Daniel T. Citron, Carlos A. Guerra, Andrew J. Dolgert, Sean L. Wu, John M. Henry, Héctor M. Sánchez C, David L. Smith

**Affiliations:** University of Washington, Institute for Health Metrics and Evaluation, 2301 Fifth Ave., Suite 600, Seattle, WA 98121, USA; Medical Care Development International, 8401 Colesville Road, Suite 425, Silver Spring, MD 20910, USA; University of California, Berkeley, Divisions of Biostatistics & Epidemiology, 2121 Berkeley Way, Berkeley, CA 94720, USA

**Author notes:** To whom correspondence should be addressed. Mailing address: Institute for Health Metrics and Evaluation, 2301 Fifth Ave., Suite 600, Seattle, WA 98121, USA. DT Citron developed the project and carried out the analysis. DL Smith provided guidance and context for the project. CA Guerra provided the data for building the Bioko Island model. SL Wu, JM Henry, HM Sánchez C, and D Dolgert discussed and reviewed the analysis. The authors declare that they have no competing interests.

**Keywords:** Mathematical epidemiology, Infectious disease modeling, Human population movement, Malaria

## Abstract

Newly available data sets present exciting opportunities to investigate how human population movement contributes to the spread of infectious diseases across large geographical distances. It is now possible to construct realistic models of infectious disease dynamics for the purposes of understanding global-scale epidemics. Nevertheless, a remaining unanswered question is how best to leverage the new data to parameterize models of movement, and whether one’s choice of movement model impacts modeled disease outcomes. We adapt three well-studied models of infectious disease dynamics, the SIR model; the SIS model; and the Ross-Macdonald model, to incorporate either of two candidate movement models. We describe the effect that the choice of movement model has on each disease model’s results, finding that in all cases there are parameter regimes where choosing one movement model instead of another has a profound impact on epidemiological outcomes. We further demonstrate the importance of choosing an appropriate movement model using the applied case of malaria transmission and importation on Bioko Island, Equatorial Guinea, finding that one model produces intelligible predictions of *R*_0_ while the other produces nonsensical results.

**Significance Statement:** Newly available large-scale datasets of human population movement represent an opportunity to model how diseases spread between different locations. Combining infectious disease models with mechanistic models of host movement enables studies of how movement drives disease transmission and importation. Here we explore in what ways modeled epidemiological outcomes may be sensitive to the modeler’s choice of movement model structure. We use three different mathematical models of disease transmission to show how a model’s epidemiological predictions can change dramatically depending on the chosen host movement model. We find these different outcomes are robust to using the same data sources to parameterize each candidate model, which we illustrate using an example of real-world malaria transmission and importation in Bioko Island, Equatorial Guinea.

---

**M**athematical models are important tools for understanding disease transmission, making it possible to estimate the size, timing, and impact of epidemics and the effectiveness of interventions. There have been many such mathematical models adapted and applied to study epidemics that spread across large geographical regions (1, 2). Many such models are often motivated by recent pandemics, such as SARS (3); Ebola virus disease (4); Zika fever (5); and the 2019 novel coronavirus (6). In each of these cases, long-distance travel of infected human hosts proved to be an important driver of the spread of infectious disease between geographically separated populations. Real world studies of the spatial dynamics of human infectious diseases must choose an underlying modeling framework for simulating human mobility which is then parametrized with mobility data and coupled with a model of disease dynamics. Here, we have conducted a study that focuses on the consequences which the choice of movement modeling framework can have on various disease dynamical processes.

Parameterizing realistic models of disease transmission occurring across wide geographical ranges is now possible because of the recent availability of large, highly detailed data sets describing human movement patterns (7). Census data attempts to describe population migration that occurs between census years (8). Traditional surveys of commuters (9, 10), patients (11, 12), or residents in disease-affected areas (13, 14) provide another description of recent travel activity. More recently, mobile phone service providers have shared privacy-protected data sets, showing how large numbers of users tend to move between cell phone towers (12, 15–18). GPS trip loggers have also recently been used to track study participants’ movement activities (19).

The recent abundance of movement data provide an exciting new opportunity to use models to quantify how human host movement affects epidemiological outcomes. Nevertheless, each method of data collected only provides a partial picture of true movement patterns, and for many studies it is necessary to build a model of host movement to fill gaps in the data or to synthesize data coming from many sources. It remains an unanswered question how best to use the data to build and parameterize models representing host movement.

How should one choose to represent movement rates based on available data? An important consideration is finding a statistical model that fits adequately well to movement data, accurately predicting the frequency of travel or flow volume between two locations. There are already many candidate movement models, such as gravity and radiation models, which have been evaluated against data in a variety of different settings (13, 18, 20–22). A fitted movement model, once it has been evaluated as sufficiently accurate, may then be used to set the movement-related parameters of a mechanistic disease transmission model.

Criteria for selecting a model to parameterize and whether the choice of model would affect the conclusions of a study has received much less attention (9, 23). For the purposes of modeling movement to understand infectious disease dynamics, the structure of the movement model—the rules which govern how hosts’ movement patterns are represented—affect the model’s quantitative behavior and subsequent predicted epidemiological outcomes. The same travel data sets used to calibrate different movement models can have different disease dynamics as a consequence of the movement model choice (9). One’s choice of movement model structure defines the set of parameters which require fitting and remains distinct from the question of how best to fit those parameters.

There is an important structural difference between two of the simplest and most commonly used classes of movement models. Eulerian movement models specify the rates at which hosts from one location travel to any other location. Eulerian models do not track individual behavior, such that after a host moves it has no memory of its previous location. Lagrangian movement models, by way of contrast, follow individuals. The class of Lagrangian movement models includes any model that specifies how frequently hosts travel away from home before returning (23, 24). The authors of (24) formulate models of vector-borne disease with Eulerian and Lagrangian host movement but they do not directly compare the models’ quantitative behavior, inviting further questions of when one host movement model might be more appropriate than another. Depending on the level of detail contained in the available data, one might be able to use the data to select a class of candidate mechanistic movement models, but the context for a problem also provides some useful information about which model might be more appropriate (23). Modelers whose aim is to understand how movement influences epidemiological outcomes must choose one such mechanistic description to represent movement.

More generally, there has never been a thorough exploration of how the modeler’s choice of movement model may influence modeled epidemiological outcomes. Most early discussions of metapopulation modeling (e.g. (25)) primarily focused on formulating the models, without discussing how one might make use of travel data for movement model calibration or how to select a movement model depending on epidemiological context. More recently, the authors of (26) offer the heuristic advice that an Eulerian movement model is suited to describing animal migration while a Lagrangian movement model is more suited to describing human commuting behavior, but they do not provide supporting quantitative analysis. At least one study has used the same travel data sets to calibrate different movement models only to find that the disease dynamics can change as a consequence of the movement model choice (9). In some cases, it may be possible to derive and compare simpler models of disease transmission by carefully rescaling movement parameters to capture the essential features of spatial dynamics in a generalized metapopulation (27). Generally speaking, however, there has not yet been a thorough quantitative investigation into whether the modeler is free to substitute one model for another for a given epidemiological setting without affecting the model’s results and conclusions.

When it comes to incorporating movement data into an epidemiological model, the problem becomes whether choosing one mechanistic movement model over another can impact modeled epidemiological outcomes, and whether there are certain epidemiological settings where one type of movement model may be more appropriate or accurate. In the present analysis we explore this problem using compartmental metapopulation models which integrate together host movement and infectious disease dynamics. We use three infectious disease transmission models—the Susceptible-Infected-Recovered model (SIR); the Susceptible-Infected-Susceptible model (SIS); and the Ross-Macdonald model—which represent a suite of tools for modeling the transmission dynamics of a wide variety of pathogens. We adapt each of the disease transmission models to incorporate two mechanistic representations of movement: the first is an Eulerian movement model which we call the Flux model, and the second is a Lagrangian movement model which we call the Simple Trip model. We directly compare the Flux and Simple Trip models by setting parameters such that the total flux of travelers between each sub-population remains constant, thus emulating a case where one might have a single data set from which one could calibrate either movement model. We examine how disease model outcomes can change with different movement models for each transmission model. For all three transmission models, we find that the modeled quantities of interest relating to disease dynamics can differ dramatically depending on one’s choice of movement model. We conclude by applying this analysis to a model of malaria transmission and importation on Bioko Island in Equatorial Guinea, and use intuition based on our prior analysis to frame our understanding for why the Flux model surprisingly fails to produce meaningful predictions of transmission intensity in that context.

## Methods

### Host movement models

We use metapopulation models to represent a network of geographically isolated populations of hosts. Each population occupies a location where local conditions may affect transmission intensity. Each population *i* contains *N*_*i*_ hosts. We assume that the population at each location remains stable over time — in applied contexts each population may be calibrated using census or other population data. Disease transmission is assumed to be completely local, such that hosts from different sub-populations come into contact with one another only if they travel to occupy the same location. We describe and compare two simple models of host movement, each of which represents a different set of rules governing how hosts move from one location to another.

The “Flux model” is an Eulerian movement model which describes hosts as diffusing from one metapopulation to another (24):

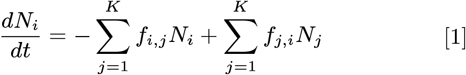

where *N*_*i*_ counts the number of hosts currently located at site *i*. The total number of hosts remains constant over time 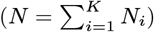. The constant *f*_*i,j*_ represents the rate at which hosts located at *i* travel to *j*, where *f*_*i,i*_ = 0 for all *i*. The fully specified Flux model requires *K*(*K* − 1) parameters.

The “Simple Trip model” is a Lagrangian movement model which assigns home locations to each host and describes how hosts travel temporarily to other locations before returning home (23, 24). Unlike the Flux model, the Simple Trip model differentiates between the residents and the visitors currently located at the same site. Visitors are able to interact with residents at a given site, but they return home at fixed rates:

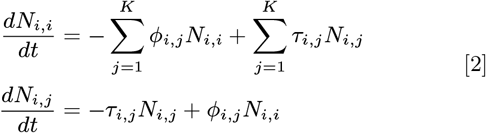

The notation in Eq. 2 has changed to account for hosts retaining memory of their home sites: a host from *i* who is currently located at *j* will be counted as belonging to the *N*_*i,j*_ population, and the number of hosts whose home is *I* remains constant over time, even if members visit other sites 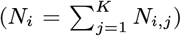. The constant *ϕ*_*i,j*_ represents the rate at which hosts whose home is *i* travel to *j*, while the constant *τ*_*i,j*_ is the rate at which hosts visiting *j* from *i* return home to *i*. Both *ϕ*_*i,i*_ = 0 and *τ*_*i,i*_ = 0 for all *i*. The fully specified Simple Trip model requires 2*K*(*K* − 1) parameters

When the movement equations reach a steady state and the derivatives on the left hand side of Eq. 2 equal zero, the population *N*_*i*_ is distributed across the *K* metapopulation sites as follows (23):

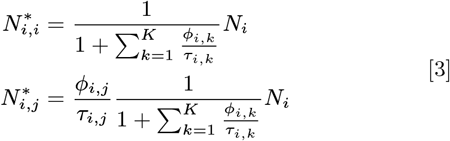

We can use Eqs. 3 to define a “Time at Risk Matrix” *ψ* where *ψ*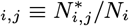, the average number of individuals from *i* found at site *j* in the steady state, or the fraction of time on average that hosts who live in site *i* spend in site *j*.

To directly compare the quantitative behavior of both models, we will assume that in the steady state the total number of hosts moving from one site to another is the same across both models. That is to say, the number of people traveling *i* → *j* in the Flux model is the same as the number of *i* residents traveling to *j* + the number of *j* resident travelers returning home from *i* in the Simple Trip model:

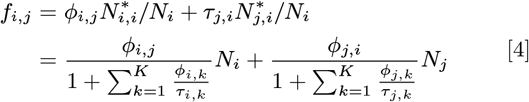

It can be shown under these circumstances that the steady state populations in the Flux model 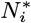 equal the stable populations in the Simple Trip model. Another property of Eq. 4 is that when we constrain the Flux model parameters to match the fluxes in the Simple Trip model, the Flux model parameters become symmetric *f*_*i,j*_ = *f*_*j,i*_. Under this condition the fully specified Flux Model only requires *K*(*K* − 1)*/*2 parameters, meaning that more detailed information is required to parameterize the Simple Trip model than the Flux model.

### SIR Models

The next step is to incorporate both movement models into three compartmental models of disease transmission. The first such model is the SIR model, which describes a single outbreak of disease. For a single population, the deterministic ordinary differential equation SIR model is as follows:

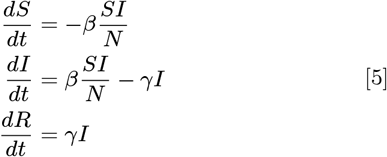

Combining the basic SIR model with the Flux movement model 1, we obtain an analogous set of 3*K* equations (23):

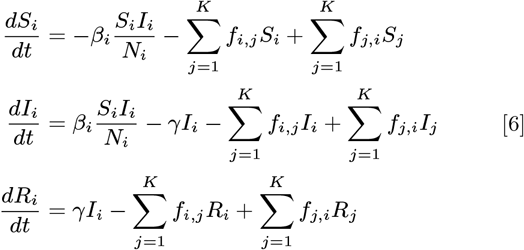

The parameters *β* and *γ* in Eq. 6 represent the transmission rate and recovery rate, respectively.

Combining the basic SIR model with the Simple Trip model 2, we obtain an analogous set of 3*K*^2^ equations describing transmission between hosts while they are at their home location or while they are traveling:

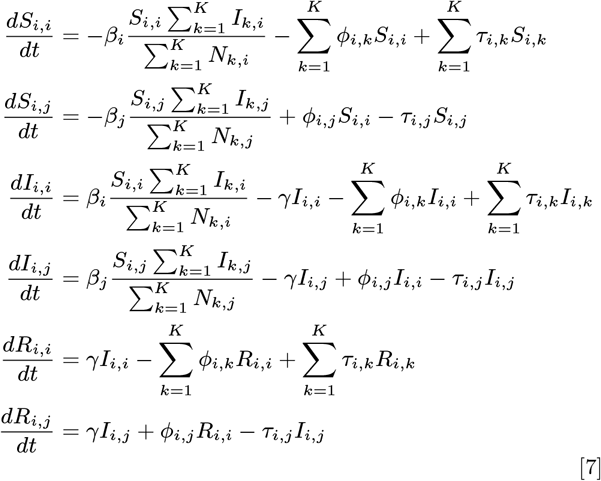

Note that for both movement models we allow the parameter representing transmission intensity *β*_*i*_ to vary across locations, not across individuals, representing how transmission intensity reflects the contact network structure or other local conditions affecting transmission. *γ*, by contrast, is held constant for all individuals regardless of location.

### SIS Models

Our second disease transmission model is the SIS model, which can be used to describe persistent endemic disease brought about by cycles of repeated infection. We adapt the SIS model to include either the Flux or Simple Trip models to describe movement between sub-populations. For a single population, the SIS model equation is as follows:

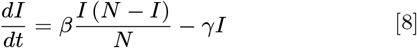

Combining the basic SIS model with the Flux model (Eq. 1), we obtain a set of *K* equations for transmission in the *i*th site:

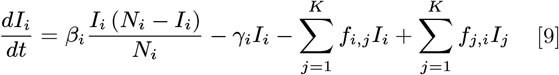

Combining the basic SIS model with the Simple Trip model (Eq. 2), we obtain a set of *K*^2^ equations among both residents at home and travelers who are away:

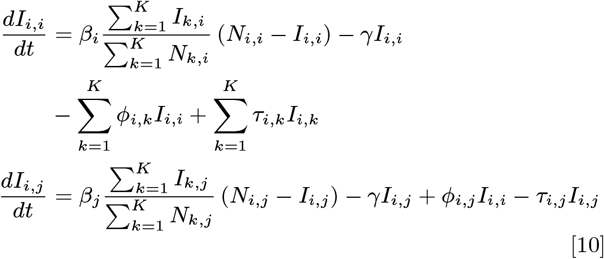

Again, in both movement models the transmission intensity *β*_*i*_ varies across locations, not across individuals, and *γ* is held constant for all individuals in all locations.

### Ross-Macdonald Models

Our third disease model is the Ross-Macdonald model, which was developed to describe malaria transmission (28, 29). The Ross-Macdonald model is similar to the SIS model, except it includes a vector population which serves as a mechanism of transmission from infected to susceptible human hosts. The Ross-Macdonald model tracks infection dynamics among human hosts (*X*) and vectors (*Z*) and depends on bionomic parameters (*M, a, b, c, g, n*, see Supplementary Information section 3 for definitions) which reflect the environment’s support of the vectors which transmit disease.

The basic Ross-Macdonald model is as follows:

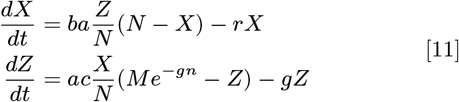

The first of Eqs. 11 describes how human hosts become infected through contact with infectious mosquitoes, while the second describes how mosquitoes become infected through contact with infectious humans. Setting the derivatives in Eq. 11 to zero represents an endemic equilibrium in which the local environment may support endemic malaria under the right conditions. Standard fixed-point analysis of the endemic equilibrium yields an expression for *R*_0_ as a function of bionomic parameters. As long as the endemic prevalence *X*^*^ ≥ 0 it is also possible to re-formulate *R*_0_ as a function of prevalence (28, 29):

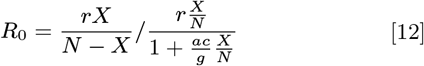

As before, we adapt the basic Ross-Macdonald model by allowing human hosts to move from one metapopulation to another. At present we will hold the parameters (*r, a, b, c, g, n*) to be constant across all sites, while the stable mosquito population supported by the local environment (*M*_*i*_) is allowed to vary across all sites. Incorporating the Flux model into the metapopulation Ross-Macdonald model (24):

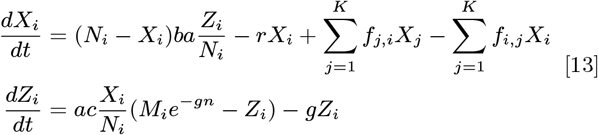

We again perform a standard analysis of the endemic equilibrium of Eqs. 13 and obtain an expression for the 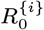 in each site as a function of prevalence at each site *X*_*i*_:

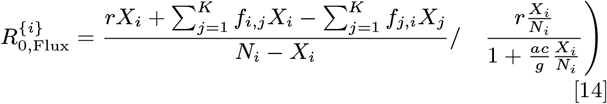

We also extend the basic Ross-Macdonald model by allowing human hosts to move from one metapopulation to another as described by the Simple Trip model. Following the analysis of (24) and (17, 30), we simplify the analysis of the equilibrium behavior of the metapopulation Ross-Macdonald model using the Time at Risk matrix *ψ*. We rewrite the force of infection experienced by human hosts as a weighted average which combines local transmission and the average fraction time spent at risk in different locations:

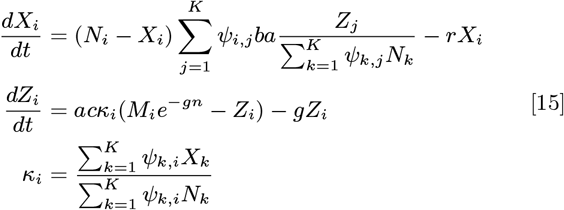

The expression *κ*_*i*_ represents the fraction of infectious humans located at *i*, accounting for both local residents as well as visitors from other locations. From the endemic equilibrium of Eq. 15, and following the analysis of (17), we obtain an expression for 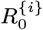 which reflects how the movement model alters transmission at each metapopulation site:

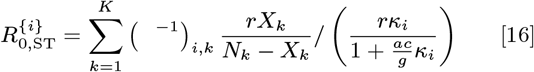

## Results

### SIR models

We begin by directly comparing the predicted outcomes of the two versions of the SIR model with Flux and Simple Trip movement. We simulate epidemics in two coupled metapopulations of *N*_*i*_ = 500 human hosts by numerically integrating Eq. 6 and Eq. 7 and measuring the size of the outbreak. The population of susceptibles remaining after the outbreak has ended *S*_∞_ quantifies the size of the outbreak and allows one to infer *R*_0_ (26). Furthermore, *S*_∞_ quantifies the population at risk in subsequent future epidemics, which is relevant for modeling seasonally recurring pathogens such as influenza (31).

We set *γ* = 1 in all locations, hold *β*_2_ constant, and measure the residual population *S*_∞_ in location 1 as *β*_1_ varies. We identify two transmission parameter regimes: *β*_2_ *> γ*, which is high enough to cause an outbreak in location 2, and *β*_2_ *< γ*, which in isolation does not lead to an outbreak in location 2. We also define two travel parameter regimes, one where travel occurs frequently and lasts short periods of time (*τ*_*i,j*_ *> γ*), and one where travel occurs infrequently and lasts long periods of time (*τ*_*i,j*_ *< γ*). We specify the parameter values for *ϕ*_*i,j*_ and *τ*_*i,j*_ in each of these regimes for the Simple Trip model and use Eq. 4 to also parameterize the Flux model, such that we emulate using the same data inputs to parameterize both movement models.

Figure 1 compares how the two SIR models with metapopulation movement behave in different parameter regimes. In all cases the outcomes are dramatically different, showing that SIR model predictions depend strongly on how host movement is represented in the metapopulation model. The two movement models do agree when transmission parameters are equal in both locations (*β*_1_ = *β*_2_), but there are large quantitative differences whenever there is some heterogeneity in the local transmission conditions across the different locations.

**Fig. 1.**
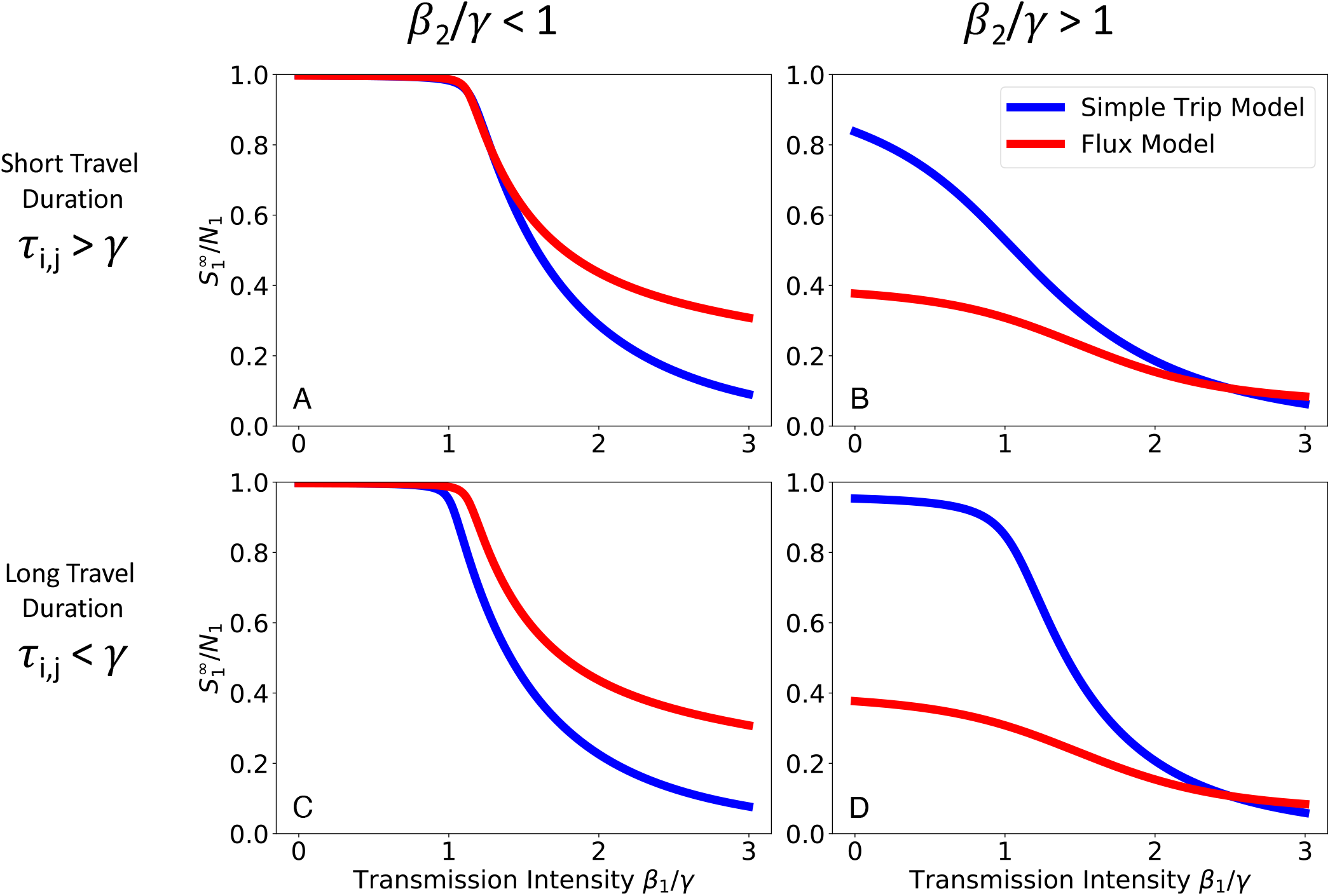
Comparing SIR model results. We numerically integrate Eqs. 6 and 7 to find the size of the residual population of susceptibles following the outbreak. We plot the residual population fraction in location 1 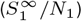 as a function of transmission intensity in location 1 (*β* _1_ / γ) while holding transmission intensity in location 2 (*β* _2_ / *γ*) constant. We explore four different parameter regimes defined by the transmission intensity of location 2 and the duration of travel. In all four parameter regimes there is a dramatic separation between the predicted outbreak sizes that depends on whether one uses the Flux (red) or the Simple Trip (blue) model. As long as residents of location 1 travel from a low to a high transmission environment (*β* _2_ > *β* _1_) the Simple Trip model’s outbreak will be smaller and the residual population will be larger; the opposite is true when residents of location 1 travel from a high to a low transmission environment (*β* _2_ < *β* _1_). **A**: Short travel duration regime (ϕ _i_ = 1, *τ* _i, j_ = 10); low transmission in location 2 (*β* _2_ = 0. 5) **B**: Short travel duration regime (ϕ _i_ = 1, *τ* _i, j_ = 10); high transmission in location 2 (*β* _2_ = 2. 5) **C**: Long travel duration regime (ϕ_i_ = 0. 01, *τ* _i, j_ = 0. 1); low transmission in location 2 (*β* _2_ = 0 .5) **D**: Long travel duration regime (ϕ _i_ = 0 .01, *τ* _i, j_ = 0 .1); high transmission in location 2 (*β* _2_ = 2 .5)

The major difference between the Flux and Simple Trip models is that the Simple Trip model constrains the amount of time that a traveler spends away. For example, the larger residual population for the Simple Trip model, seen in the right hand column of Figure 1, follows from a limit on the amount of time that residents of population one spend in the high-transmission location 2. Another difference between the Flux and the Simple Trip models is that hosts in the Flux model have no home residence and continue to move freely between populations after the outbreak ends. As a result, as the epidemic dies out the fractions of susceptibles become constant across all locations. In contrast with the Simple Trip model, the Flux model effectively erases any evidence that the transmission intensity differed between the two locations.

### SIS models

We next compare the respective behavior of the SIS Flux metapopulation model (Eq. 9) and the SIS Simple Trip metapopulation model (Eq. 10) for *K* = 2 metapopulations of *N*_*i*_ = 500 coupled together. Again, we set *γ* = 1 at all locations. The outcome of interest for the SIS model is now prevalence, which we find by numerically solving for the endemic equilbria in Eq. 9 and Eq. 10. We hold *β*_2_ constant and solve for the prevalence in location 1 (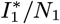 for the Flux model and 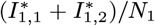 for the Simple Trip model) as a function of the transmission intensity in location 1 (*β*_1_).

We define two transmission parameter regimes: *β*_2_ *> γ*, which is high enough to sustain endemic transmission locally in location 2, and *β*_2_ *< γ*, which cannot locally sustain endemic transmission. We again define two travel parameter regimes, one where travel occurs frequently and over short periods of time (*τ*_*i,j*_ *> γ*) and one where travel occurs infrequently and over long periods of time (*τ*_*i,j*_ *< γ*). In each regime we specify travel rate parameter values for the Simple Trip model and use Eq. 4 to set parameters for the Flux model.

Figure 2 shows the results from both the Simple Trip and Flux models in each of the four parameter regimes. In the short travel duration regime (2 A, B), as long as the transmission intensity differs between the two locations, there is a noticeable separation between the two movement models’ predicted relationships between prevalence and local transmission intensity. In 2 A it is even the case that the apparent critical point, above which location 1 has sustained endemic disease, changes depending on which movement model is used.

**Fig. 2.**
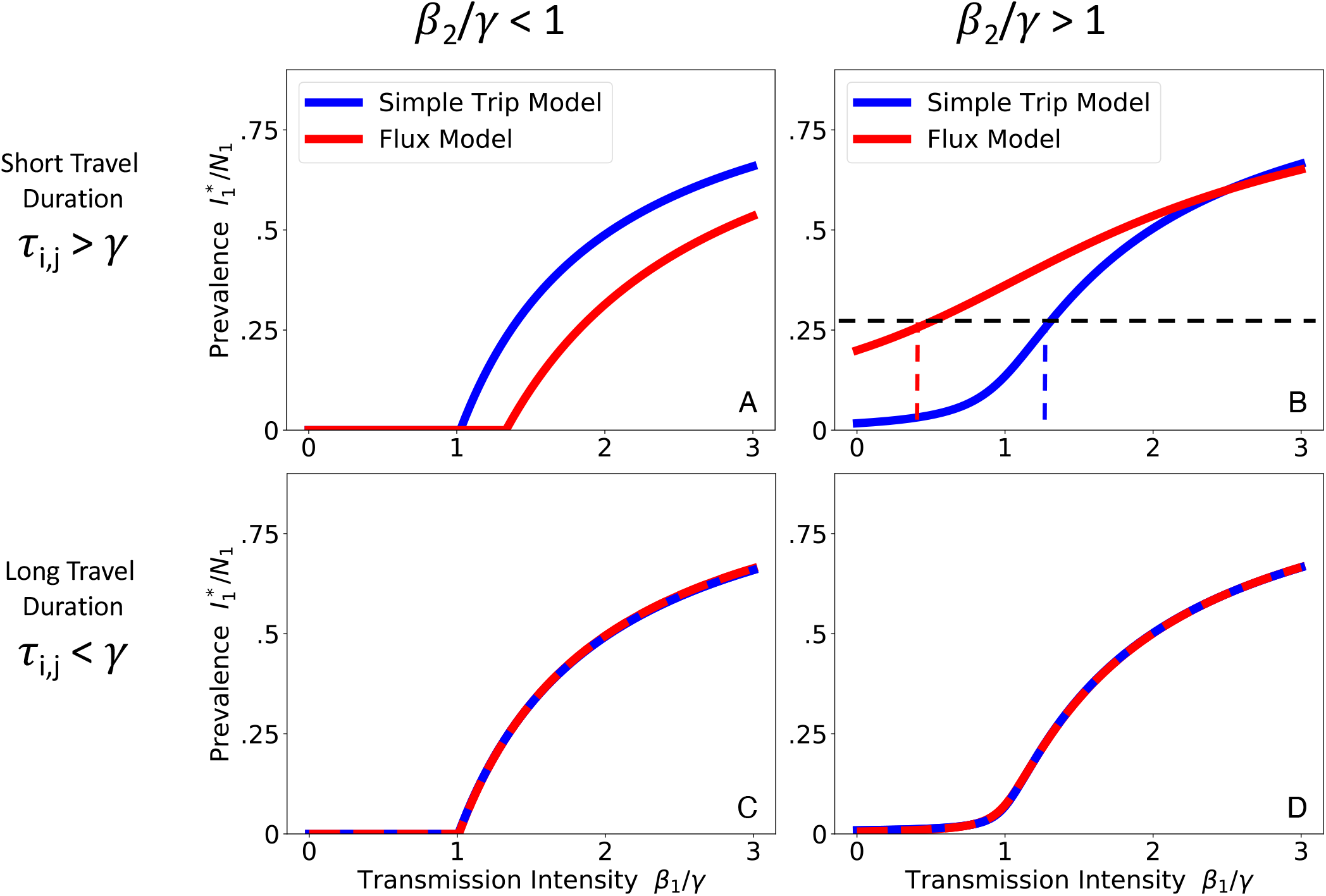
Comparing SIS model results. We numerically solve Eqs. 9 and 10 for the prevalence at endemic equilibrium. We plot the endemic prevalence in location 1 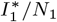 as a function of transmission intensity in location 1 (*β*_1_ */γ*) while holding transmission intensity in location 2 (*β*_2_ */γ*) constant. We explore four different parameter regimes defined by the transmission intensity of location 2 and the duration of travel. When travel duration is short (upper row), the Flux (red) and Simple Trip (blue) models produce dramatically different relationships between transmission intensity and prevalence. When travel duration is long (lower row), the two movement models’ results converge. **A**: Short travel duration regime (*ϕ*_*i*_ = 0.5, *τ*_*i,j*_ = 40); low transmission in location 2 (*β*_2_ = 0.5). **B**: Short travel duration regime (*ϕ*_*i*_ = 0.5, *τ*_*i,j*_ = 40); high transmission in location 2 (*β*_2_ = 2.5). **C**: Long travel duration regime, 9*ϕ*_*i*_ = 0.005, *τ*_*i,j*_ = 0.4); low transmission in location 2 (*β*_2_ = 0.5). **D**: Long travel duration regime, 9*ϕ*_*i*_ = 0.005, *τ*_*i,j*_ = 0.4); high transmission in location 2 (*β*_2_ = 2.5).

The discrepancy in the short travel duration regime arises because the Simple Trip model imposes an extra constraint on the amount of time a traveler spends away from home. The Flux model does not specify the amount of time spent away, and its movement rates (from Eq. 4) tend to overestimate the time spent away. The Simple Trip model endemic prevalence in 2 A is elevated because travelers spend less of their time at risk in the low transmission location 2. Similarly, the Simple Trip model endemic prevalence in Figure 2 B is suppressed because the amount of time that travelers from location 1 spend at risk in the high transmission location 2 is constrained to be smaller. In the long travel duration regime (Figure 2, lower row), however, if the trip time is longer than the disease infectious period there is enough time for prevalence among travelers to become equal to the prevalence among residents of location 2: in this parameter regime, the Flux model and Simple trip models converge, because the constraint on time spent away from home makes less of a difference.

### Ross-Macdonald models

We examine the behavior of the Ross-Macdonald models with Flux and Simple Trip movement. We use the model to represent a malaria-causing pathogen with a mean infectious period of *r*^−1^ = 200 days. The outcome of interest in the Ross-Macdonald model is *R*_0_, which quantifies the local transmission intensity required to sustain endemic malaria at observed prevalence levels. We compare the two movement models using *K* = 2 connected sub-populations of *N*_*i*_ = 500 human hosts by specifying prevalence and using Eqs. 14 and 16 to calculate *R*_0_ for the Flux and Simple Trip models, respectively. Again, we define two parameter regimes based on high (*X* = 0.25) versus low (*X* = 0.025) prevalence in location 2, and another pair of parameter regimes based on high frequency, short duration trips (*ϕ*_*i*_ = 1*/*360, *τ*_*i,j*_ = 1*/*10) versus low frequency, long duration trips (*ϕ*_*i*_ = 1*/*72000, *τ*_*i,j*_ = 1*/*4000). As before, Flux model parameters are defined from the Simple Trip model parameters using Eq. 4.

Figure 3 illustrates how prevalence in location 1 varies in relation to *R*_0_ in location 1. As long as the prevalences in each location are different, there is a dramatic separation between the Flux and Simple Trip model results, particularly in the short travel duration regime (Figure 3 A, B). If travel duration is long, however, the two models’ predicted relationships between prevalence and *R*_0_ do approach one another. The relationships between prevalence and transmission intensity for the two Ross-Macdonald models are qualitatively similar to the one shown previously for the SIS model in Figure 2.

**Fig. 3.**
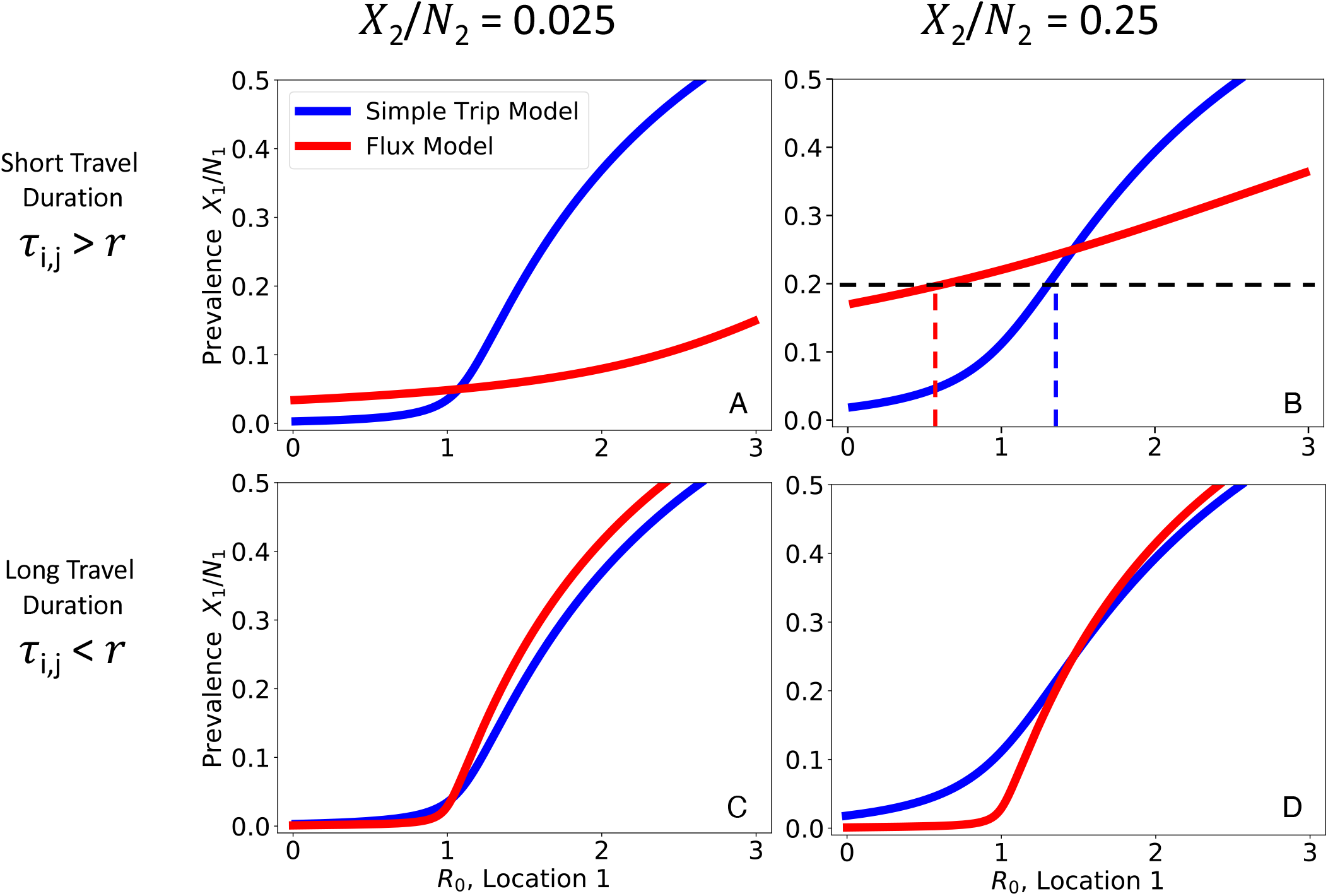
Comparing Ross-Macdonald model results. We numerically solve Eqs. 14 and 16 to find the relationship between prevalence in location 1 (*X*_1_ */N*_1_) and transmission intensity (*R*_0_). We plot the relationship between location 1 prevalence (*X*_1_ */N*_1_) as a function of transmission intensity in location 1 while holding prevalence constant (*X*_2_ */N*_2_) in location 2. Note that we plot prevalence on the y-axis and transmission intensity on the x-axis, to emphasize how the Ross-Macdonald model behaves similarly to the SIS model (Figure 2). We explore four different parameter regimes defined by the prevalence of location 2 and the duration of travel in relation to the average duration of infection (*r*^−1^ = 200 days). In the short travel duration regime, there is a large quantitative difference in the relationship between prevalence and *R*_0_ for the two different movement models. In the regime with short travel duration and high location 2 prevalence (upper right), for a given prevalence of 0.2 using the Flux model would suggest low transmission 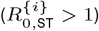, whereas using the Simple Trip model would suggest high transmission 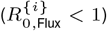. In the long travel duration regime, the quantitative disagreement between the two movement models is less significant. **A**: Short travel duration regime (*ϕ*_*i*_ = 1*/*180, *τ*_*i,j*_ = 1*/*10); low prevalence in location 2 (*X*_2_ */N*_2_ = 0.025) **B**: Short travel duration regime (*ϕ*_*i*_ = 1*/*180, *τ*_*i,j*_ = 1*/*10); high prevalence in location 2 (*X*_2_ */N*_2_ = 0.25) **C**: Long travel duration regime (*ϕ*_*i*_ = 1*/*72000, *τ*_*i,j*_ = 1*/*4000); low prevalence in location 2 (*X*_2_ */N*_2_ = 0.025) **D**: Long travel duration regime (*ϕ*_*i*_ = 1*/*72000, *τ*_*i,j*_ = 1*/*4000); high prevalence in location 2 (*X*_2_ */N*_2_ = 0.25)

There are some ranges of prevalence that are not allowed as long as *R*_0_ *>* 0, such as the Flux model predictions in Figure 3 B. This occurs when incidence and, by consequence, prevalence among travelers to location 2 is so high that it raises the base prevalence in location 1. For example, in the Flux model shown in Figure 3 B, the relationship between transmission, movement, and prevalence specified by the model makes it impossible to reconcile high prevalence in location 2 with very low prevalence in location 1. The transmission model creates an effective lower bound on prevalence in location 1, and we interpret prevalence values below that lower bound as having no valid solution which is consistent with the transmission model.

### Malaria Modeling Example

We further illustrate the quantitative disagreements between the Flux and Simple Trip models by applying the Ross-Macdonald models with host movement to the problem of estimating local *R*_0_ (*i*.*e*. computed as if there was no host movement) in a real-world setting. Bioko Island is located in the Atlantic Ocean about 225km to the northwest of mainland Equatorial Guinea (EG) in Western Africa. From population data collected during a bednet mass-distribution campaign in 2015, human population count was estimated to be 239,000 people, about 85% of whom lived in EG’s capital city Malabo in the north (32). Following the recent efforts of an intensive malaria control and elimination program (Bioko Island Malaria Elimination Program, BIMEP), the average malaria parasite rate (PR, a measure of prevalence) in children 2-14 years old has fallen from an average of 0.43 to 0.11 between 2004 and 2016, with further progress appearing to have stagnated since 2015 (33). A recent analysis of data collected through malaria indicator surveys (MIS) (34–36) has used geostatistical techniques (37) to map the estimated prevalence of malaria (Figure 4 A) and the estimated probability of a resident leaving (Figure 4 B) across different locations on Bioko Island (14). The analysis showed an elevated risk of infection among people who reported traveling recently (14), a pattern corroborated by other studies (33, 38). In particular, this suggests that travelers who had spent time in mainland EG, which still has an estimated mean prevalence of around 0.43 (39, 40), are very likely to bring back infections with them that would contribute to the sustained endemic prevalence measured on Bioko Island.

**Fig. 4.**
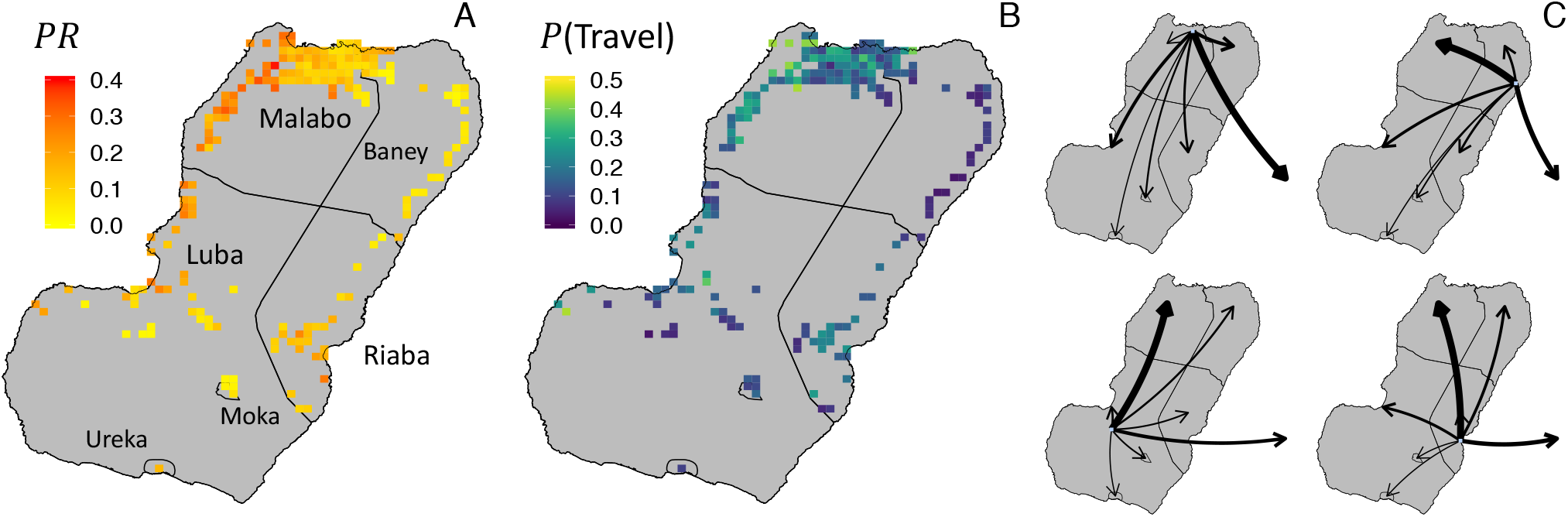
Parameterizing Ross-Macdonald model from data. **A** Prevalence (Parasite Rate, *X*_*i*_*/N*_*i*_) map, estimated from the MIS collected parasitemia measurements, reproduced with permission from (14). Each pixel shows the mean estimated prevalence within a 1 *×* 1 km^2^ area. **B** Travel “prevalence” map, reproduced with permission from (14), where each pixel shows the probability that a resident leaves home during an eight week survey period. **C** Modeled rates of travel *{ϕ*_*i,k*_*}* for each of four example pixels, estimated from MIS data reporting recent travel. Arrows pointing off-island represent travel to Mainland EG. Arrow weights are proportional to rate of travel from origin pixel to each of the 7 destination regions, or the sum total rate of travel to all of the destination pixels within each region.

We can use the Ross-Macdonald model to infer local transmission intensity, described by the local *R*_0_, based on the known prevalence and travel behavior. We treat each of the pixels shown in the maps in Figure 4 as its own sub-population with its own local transmission environment, which allows the model to contain the spatial heterogeneity known from the data. We use the population data to set the population *N*_*i*_ and the geospatial estimates to set the prevalence *X*_*i*_*/N*_*i*_ at each location. For the travel parameters we use trip destinations reported in the MIS (41) to fit a multinomial probability model of destination choice. Together with the travel frequency data (Figure 4 B), we find a set of travel frequency parameters {*ϕ*_*i,k*_}, illustrated in Figure 4 C. We use MIS data on trip duration (41) to fit an exponential model of trip duration and find that travel within Bioko Island and to mainland equatorial had a mean duration of 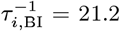 and 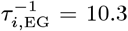 days, respectively. (Further details may be found in Supplementary Information section 2.) The rate parameters *f*_*i,j*_ for the Flux model were calculated using Eq. 4.

In this way, we construct a fully parameterized pair of Ross-Macdonald models with movement between sub-populations, from which we can infer the transmission intensity in each location on Bioko Island. Figure 5 A maps estimates of *R*_0,Flux_ calculated using the Flux model (Eq. 14). The dark pixels of the map represent areas where there is no valid solution for *R*_0,Flux_. In these locations, the rate at which people become infected while traveling is too high for the transmission model to reconcile local transmission intensity with local prevalence and the volume of imported infections. Figure 5 B maps estimates of *R*_0,ST_ calculated using the Simple Trip model (Eq. 16). In contrast with the Flux model, the *R*_0,ST_ estimates are easier to interpret. Local transmission *R*_0,ST_ ≥ 1 along the northwest coast and around Riaba. *R*_0,ST_ ≤ 1 in many places in Malabo, suggesting that the sustained prevalence in this urban area may be attributable to travelers returning from the mainland with imported infections. Figure 5 C compares the distributions of *R*_0,Flux_ and *R*_0,ST_, further contrasting the two sets of predictions and further illustrating how the Flux model fails to produce meaningful results in some areas.

**Fig. 5.**
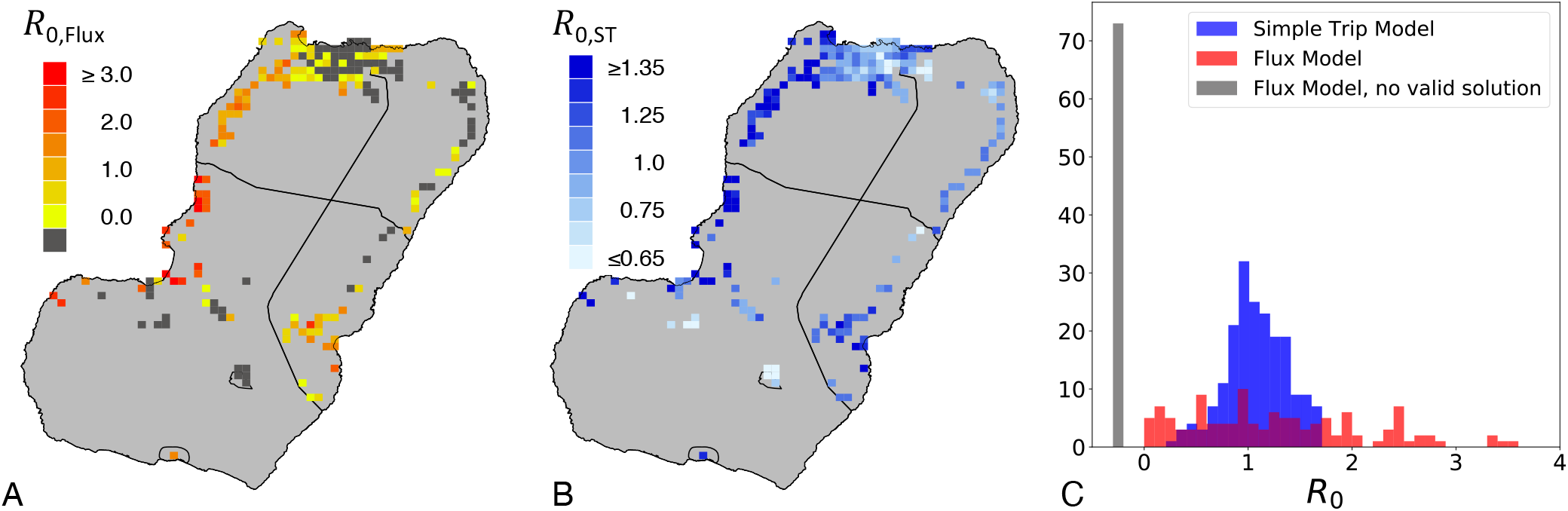
Modeled estimates of *R*_0_. All modeled estimates were generated using the same prevalence and travel data, illustrated in Figure 4. **A**: *R*_0,Flux_, calculated using the Flux model (Eq. 14) parameterized with the input data shown in Figure 4. Dark gray denotes areas where there is no valid solution for *R*_0,Flux_, meaning that according to the Flux model travelers are subject to too high transmission risk when on the mainland. **B**: *R*_0,ST_, calculated using the Simple Trip model (Eq. 16) parameterized with the input data shown in Figure 4. **C**: Histograms comparing the distributions of *R*_0,Flux_ and *R*_0,ST_ calculated across the island, with the inset showing the full range of the Flux model results. The dark gray column of values counts the areas where *R*_0,Flux_ has no valid solution.

We frame our understanding of these results in terms of the *K* = 2 sub-population example discussed previously for the Ross-Macdonald Model. Each sub-population on Bioko Island exists in a parameter regime where travel duration is short (*τ*_*i,j*_ *> r*) and the prevalence in the off-island travel destination is higher than the local prevalence. We recall the results shown in Figure 3 A, where we see how for low prevalence values the Flux model tends to underestimate *R*_0_ compared to the Simple Trip model. The Simple Trip model constrains the amount of time that travelers spend on mainland EG, and therefore reduces the number of people who return home from traveling with malaria infections. The Flux model does not include such a constraint, and in this parameter regime we see how in some locations the rate of imported infections is so high that the model breaks down and does not allow meaningful values of *R*_0,Flux_.

## Discussion

We have shown that models of infectious disease dynamics are sensitive to the choices that modelers make when including movement-mediated interactions between sub-populations. To do so, we have adapted each of three transmission models to include two types of movement between sub-populations. For all three transmission models we have identified parameter regimes where choosing to incorporate one movement model instead of the other will result in dramatically different results. Specifically, when there is a difference in transmission intensity between two interacting locations, and when mean travel duration is short compared to the mean duration of infection.

In all instances we set movement parameters to match the total flux of individuals moving between each pair of sub-populations. In this way, we have emulated a situation where the modeler has a single movement data set and must choose which movement model to implement. The full impact of the choice of movement model became clear from the example of modeling malaria transmission and importation on Bioko Island: We calibrated each of the two movement models using the same data set and found that each movement model produced dramatically different results. In some areas, the Flux model and was unable to produce meaningful values of *R*_0,Flux_.

The Simple Trip model, by specifying the rate at which travelers return home, adds a constraint on the amount of time that travelers spend while away from home. As a consequence, the Simple Trip model constrains the amount of risk experienced by travelers. The Flux model has no such constraint: as long as there are variations in the transmission intensity across different locations there will be significant differences between the two movement models’ results.

The way that each of the movement models contributes to the infectious disease model results can lead to real confusion over the true relationship between transmission intensity and epidemiological outcomes. In some applications, as with the Bioko Island example, the purpose of the model is to infer transmission conditions, which may be difficult to measure empirically, based on more readily available prevalence or incidence measures. Returning to Figure 2 B, we can imagine trying to determine the transmission intensity *β*_1_ based on a measured prevalence of 0.25. From the Flux model we infer *β*_1_ *<* 1, which suggests that local transmission risk is low. From the Simple Trip model, however, we infer *β*_1_ *>* 1, which suggests the opposite. A similar inconsistency is illustrated for the Ross-Macdonald model in Figure 3 B, and appears again in the Bioko Island example in Figure 5.

In the case of modeling malaria transmission and importation on Bioko Island we have shown how the Flux model is not suitable for producing meaningful results. Despite this drawback, the Flux model remains an attractive option for creating a basic quantitative description of host movement in other epidemiological contexts. The Flux model requires fewer parameters to specify completely. For this reason, parameterization is possible with relatively simple data sets which count the number of individuals moving from one each location to each other location over time. Unfortunately, data sets which record flux volumes do not necessarily lend themselves to parameterizing Simple Trip models, as they do not distinguish between the rates of leaving and returning home.

For example, mobile phone call data records (CDRs) are a source of high-volume data sets for measuring the number of individuals who travel from one location to another. Often, for the sake of preserving subscriber privacy, CDRs are aggregated across individual users, time, and geographical area before becoming public. That is to say, publicly available CDR data sets count the number of mobile phones traveling from one service tower catchment area to another on a daily or weekly basis: these data sets do not record an individual’s rate of leaving home or time spent while traveling away from home. The loss of individual identity from data sets has already been shown to impact the predicted outcome of disease models in other contexts (9). While the limits and biases inherent to CDR data sets have been well-documented (42), the present study suggests an additional drawback when it comes to using CDR data sets for epidemiological modeling: because it is not clear that a Flux model is sufficient to produce accurate modeling results in some contexts, it is likewise not clear that flux data are sufficient to quantify travel behavior when building a movement model.

This is not to say that CDR data or Flux models are useless in every context. Our aim is that the present study serve as a starting point for understanding when the Flux model and Simple Trip model are interchangeable. In some settings, the conclusions may not be affected by the choice of a model, but one needs to carefully consider the epidemiological context before choosing a movement model. More generally, the question of which model is appropriate to use in context or how to model the spatial dynamics of pathogens merits greater attention (9, 23, 27).

The present work is restricted to discussing two basic deterministic models of movement. In theory, one may imagine similarly comparing more detailed and complicated movement models, such as incorporating travel behavior with multiple stops; adding stochastic effects; or using agent-based models to incorporate movement behavior heterogeneity among the travelers. It remains to be seen the extent to which such additional model features might further affect the epidemiological outcomes of interest. In practice, however, the data required for calibrating such models may not be readily available, and using a Flux or Simple Trip description of travel behavior may be more practical. Even for such simple movement models there is a risk of misusing the travel data that are available to calibrate a movement model which cannot be reconciled with the data and produces outputs that are difficult or impossible to interpret.

## Supporting Data Sets

Refer to section 4 of the Supplementary Information to find code and data supporting the analysis presented in the Malaria Modeling Example section and Figures 4 and 5.

## Data Availability

Data and supporting code are available at https://doi.org/10.6084/m9.figshare.12084831.v2

## ACKNOWLEDGMENTS

We acknowledge support from the Bill & Melinda Gates Foundation, Grant OPP1110495. Additionally, we thank the participants of Bioko Island who have taken part in the surveys from which the Bioko data were drawn, the BIMEP field surveyors and supervisors who collected these data, as well as the National Malaria Control Program and the Ministry of Health and Social Welfare of Equatorial Guinea, Marathon Oil, Nobel Energy and partners for their continued support in producing these data sets. Lastly, we would like to thank the following collaborators for their helpful insights and discussion: Joel C. Miller; Nick W. Ruktanonchai; M.U.G. Kraemer; Amy Wesolowski; John R. Giles; Guillermo A. García; Dianna Hergott; Su Yun Kang; Katherine E. Battle; and Daniel J. Weiss.

## References

1. V Colizza, A Barrat, M Barthelemy, A Vespignani, The role of the airline transportation network in the prediction and predictability of global epidemics. Proc. Natl. Acad. Sci. 103, 2015–2020 (2006).

2. S Riley, Large-scale spatial-transmission models of infectious disease. Science 316, 1298– 1301 (2007).

3. D Brockmann, D Helbing, The hidden geometry of complex, network-driven contagion phenomena. Science 342, 1337–1342 (2013).

4. MF Gomes, et al., Assessing the international spreading risk associated with the 2014 west african ebola outbreak. PLoS currents 6 (2014).

5. Q Zhang, et al., Spread of Zika virus in the Americas. Proc. Natl. Acad. Sci. 114, E4334– E4343 (2017).

6. M Chinazzi, et al., The effect of travel restrictions on the spread of the 2019 novel coronavirus (COVID-19) outbreak. Science 9757, 1–12 (2020).

7. DK Pindolia, et al., Human movement data for malaria control and elimination strategic plan-ning. Malar. journal 11, 1–16 (2012).

8. AJ Tatem, DL Smith, International population movements and regional Plasmodium falciparum malaria elimination strategies. Proc. Natl. Acad. Sci. 107, 12222–12227 (2010).

9. MJ Keeling, L Danon, MC Vernon, TA House, Individual identity and movement networks for disease metapopulations. Proc. Natl. Acad. Sci. 107, 8866–8870 (2010).

10. D Balcan, et al., Multiscale mobility networks and the spatial spreading of infectious diseases. Proc. Natl. Acad. Sci. 106, 21484–21489 (2009).

11. N Tejedor-Garavito, et al., Travel patterns and demographic characteristics of malaria cases in Swaziland, 2010-2014. Malar. J. 16, 1–18 (2017).

12. HH Chang, et al., Mapping imported malaria in Bangladesh using parasite genetic and human mobility data. eLife 8, e43481 (2019).

13. JM Marshall, et al., Mathematical models of human mobility of relevance to malaria transmission in Africa. Sci. Reports 8, 7713 (2018).

14. CA Guerra, et al., Human mobility patterns and malaria importation on Bioko Island. Nat. Commun. 10, 2332 (2019).

15. A Le Menach, et al., Travel risk, malaria importation and malaria transmission in Zanzibar. Sci. Reports 1, 1–7 (2011).

16. A Wesolowski, et al., Quantifying the impact of human mobility on malaria. Science 338, 267–270 (2012).

17. NW Ruktanonchai, et al., Identifying malaria transmission foci for elimination using human mobility data. PLoS Comput. Biol. 12, 1–19 (2016).

18. MUG Kraemer, et al., Utilizing general human movement models to predict the spread of emerging infectious diseases in resource poor settings. Sci. Reports 9, 5151 (2019).

19. KM Searle, et al., Characterizing and quantifying human movement patterns using GPS data loggers in an area approaching malaria elimination in rural southern Zambia. Royal Soc. Open Sci. 4, 170046 (2017).

20. F Simini, MC González, A Maritan, A. Barabási, A universal model for mobility and migration patterns. Nature 484, 96–100 (2012).

21. A Wesolowski, WP O’Meara, N Eagle, AJ Tatem, CO Buckee, Evaluating spatial interaction models for regional mobility in sub-Saharan Africa. PLoS Comput. Biol. 11, 1–16 (2015).

22. O. Bjørnstad, BT Grenfell, C Viboud, AA King, Comparison of alternative models of human movement and the spread of disease. bioRxiv (2019).

23. L Sattenspiel, K Dietz, A structured epidemic model incorporating geographic mobility among regions. Math. biosciences 128, 71–92 (1995).

24. C Cosner, et al., The effects of human movement on the persistence of vector-borne diseases. J. Theor. Biol. 258, 550–560 (2009).

25. RM Anderson, RM May, Infectious diseases of humans: dynamics and control. (Oxford university press), (1992).

26. MJ Keeling, P Rohani, Modeling infectious diseases in humans and animals. (2011).

27. A Iggidr, tet al., Vector borne diseases on an urban environment: The effects of heterogeneity and human circulation. Ecol. Complex. 30, 76–90 (2017).

28. JL Aron, RM May, The population dynamics of malaria in The population dynamics of infectious diseases: theory and applications. (Springer), pp. 139–179 (1982).

29. DL Smith, et al., Ross, Macdonald, and a theory for the dynamics and control of mosquito-transmitted pathogens. PLoS Pathog. 8 (2012).

30. NW Ruktanonchai, DL Smith, P De Leenheer, Parasite sources and sinks in a patched Ross-Macdonald malaria model with human and mosquito movement: Implications for control. Math. Biosci. 279, 90–101 (2016).

31. M Rypdal, G Sugihara, Inter-outbreak stability reflects the size of the susceptible pool and forecasts magnitudes of seasonal epidemics. Nat. Commun. 10 (2019).

32. The Government of Equatorial Guinea, Medical Care Development International (MCDI), Bioko Island Malaria Control Project (BIMCP) & Equatorial Guinea Malaria Vaccine Initiative (EGMVI) 4th Quarter Progress Report and Annual Review, (Medical Care Development International), Technical report (2015).

33. J Cook, et al., Trends in parasite prevalence following 13 years of malaria interventions on Bioko island, Equatorial Guinea: 2004-2016. Malar. J. 17, 1–13 (2018).

34. The Government of Equatorial Guinea, Medical Care Development International (MCDI), The bioko island malaria control project malaria indicator survey (MIS) 2015, (Medical Care Development International), Technical report (2015).

35. The Government of Equatorial Guinea, Medical Care Development International (MCDI), Bioko island malaria control project iii - malaria indicator survey (MIS) 2016, (Medical Care Development International), Technical report (2016).

36. The Government of Equatorial Guinea, Medical Care Development International (MCDI), The bioko island malaria control project malaria indicator survey (MIS) 2017, (Medical Care Development International), Technical report (2017).

37. SY Kang, et al., Spatio-temporal mapping of Madagascar’s Malaria Indicator Survey results to assess Plasmodium falciparum endemicity trends between 2011 and 2016. BMC Medicine 16, 1–15 (2018).

38. J Bradley, et al., Infection importation : a key challenge to malaria elimination on Bioko Island, Equatorial Guinea. Malar. J. 14, 1–7 (2015).

39. P Ncogo, et al., Malaria prevalence in Bata district, Equatorial Guinea: a cross-sectional study. Malar. J. 14, 1–10 (2015).

40. PW Gething, et al., A new world malaria map: Plasmodium falciparum endemicity in 2010. Malar. J. 10, 378 (2011).

41. CA Guerra, DT Citron, G. García, DL Smith, Characterising malaria connectivity using malaria indicator survey data. Malar. J. 18, 440 (2019).

42. A Wesolowski, CO Buckee, K Engø-Monsen, CJ Metcalf, Connecting mobility to infectious diseases: The promise and limits of mobile phone data. J. Infect. Dis. 214, S414–S420 (2016).

