## Supplementary Information for "Comparing Metapopulation Dynamics of Infectious Diseases under Different Models of Human Movement"

This manuscript was compiled on March 31, 2021

### 1. SIR Model Time Series

As a supplement to Figure 1 in the main text, Figure 1 shows example trajectories from integrating the SIR model forward in time (Eqs. 6 and 7). We set $\gamma = 1$ and $\beta_1/\gamma = 2$, and numerically integrate Eqs. 6 and 7 to generate example trajectories which show how the number of susceptibles decreases over time during an SIR outbreak. The example trajectories are shown for four different parameter regimes which depend on whether the transmission intensity of location 2 is high or low, and whether the duration of travel is short or long.

In all four parameter regimes there is a marked difference between the trajectories for the Flux model and the Simple Trip model. In the regime where $\beta_2/\gamma > 1$, it appears that the Flux model's predicted outbreak is more severe and results in more people becoming infected. Because the Simple Trip model constrains the amount of time that residents of location 1 spend in location 2, the Simple Trip model effectively limits exposure to the higher transmission intensity in location 2, resulting in a smaller outbreak.

We note that the Flux model allows recovered and susceptible individuals both to continue moving between locations after the outbreak has ended. The long-term equilibrium in the Flux model is that each location have the same proportion of susceptible individuals. It follows then that the $R_{0,\text{Flux}}$ calculated from $S^\infty$ is the same in every location. By contrast, the Simple Trip model does not allow for the same equilibration across locations, as susceptible individuals return to their homes following the outbreak. As a result, $R_{0,\text{ST}}$ calculated from $S^\infty$ can be different in each location.

One apparent consequence of the Flux model's tendency to equilibrate the proportion of residual susceptibles is the apparent "back-filling" behavior of susceptibles returning to higher-transmission locations following an outbreak. In the regime where $\beta_2/\gamma < 1$, the Flux model predicts a non-monotonic trend in the number of susceptibles. After the epidemic has finished spreading through much of the population in location 1, many more susceptible individuals remain in the lower-transmission location 2, who then may travel to location 1. The Flux model shows this behavior in parameter regimes where there are strong differences in transmission intensity between different locations. The Simple Trip model, by contrast, shows no such behavior.

### 2. Bioko Island Travel Model

In this section we discuss in greater detail the data sources used to parameterize the model of malaria transmission on Bioko Island discussed in the main text.

We use a number of different data sets collected by the Bioko Island Malaria Elimination Program to calibrate our models of malaria transmission. For populations ($\{N_i\}$) we use data collected during a bednet distribution campaign. Each of the pixels shown in the maps in Figure 4 represents an area known to be inhabited as of 2015.

We use a map of prevalence estimates ($\{X_i\}$, Figure 4 A) generated using geostatistical modeling techniques (1). In this example, the data are collected as part of three annual Malaria Indicator Surveys (MIS) in 2015-2017 (2–4). The MIS data set includes observations of parasitemia made using rapid diagnostic tests. The geostatistical modeling techniques are designed to estimate quantities such as prevalence from spatial data, smoothing out and correcting for noise and uneven sampling but still preserving spatial heterogeneity (5). The geospatial prevalence estimates for Bioko Island were first reported in (6) and are re-used here with permission.

The remaining unknown quantities required to calculate $R_0$ in different areas on Bioko Island using the Ross-Macdonald model are $\{\phi_{i,j}\}$, the rates at which people travel from their homes at $i$ to $j$, and $\{\tau_{i,j}\}$, the rates at which travelers return from $j$ to their homes at $i$. These quantities in turn may be used to calculate $\psi$ and $\{f_{i,j}\}$ as described in the text.

**A. Travel Frequency.** The MIS data set includes two questions which we can use to estimate the rates at which each person travels from their origin pixel $i$ to each other destination pixel $j$. First, respondents report whether or not they left home in the previous 60 days. Next, respondents report on traveling to one of seven possible destination regions: Malabo; Baney; Luba; Riaba; Moka; Ureka; and off-island to mainland Equatorial Guinea (labeled in Figure 4 A).

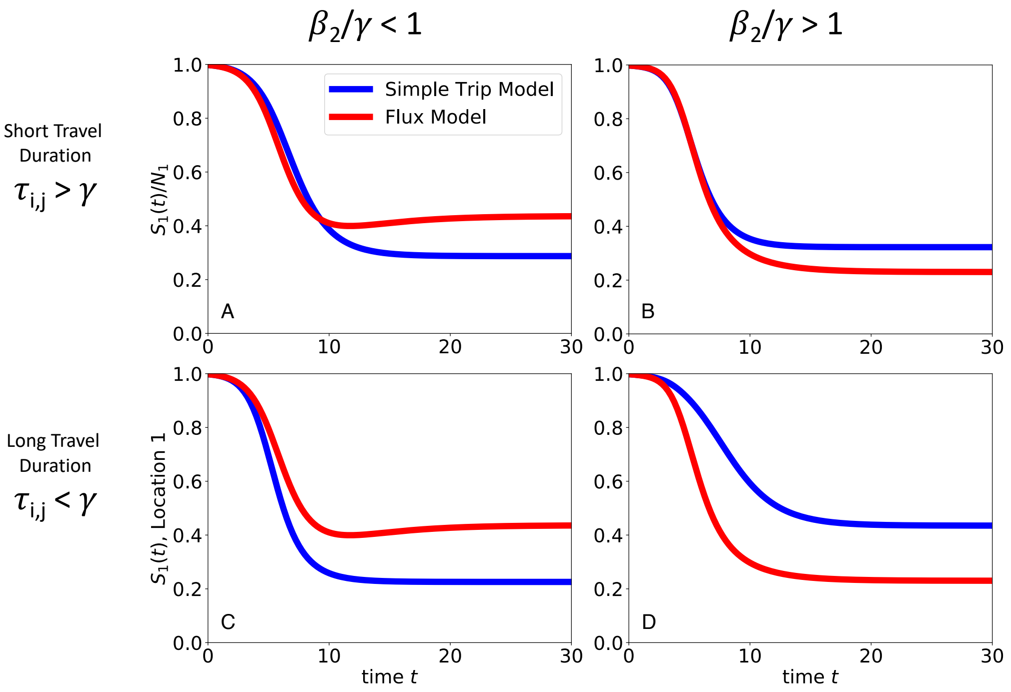


**Fig. 1. Comparing SIR model results** We numerically integrate Eqs. 6 and 7 to find example trajectories of the number of susceptibles as a function of time during an SIR outbreak. We set $\beta_1/\gamma = 2$, and explore four different parameter regimes according the transmission intensity of location 2 and the duration of travel. In all four parameter regimes, there is a marked difference between the trajectories for the Flux model (red) and the Simple Trip model (blue). In the high transmission regime ($\beta_2/\gamma > 1$), we find that the Simple Trip model's outbreak appears to end faster than the Flux model's outbreak, resulting in a larger residual susceptible population. In the low transmission regime ($\beta_1/\gamma_1 < 1$), we find that the Simple Trip model predicts a smooth sigmoidal decrease in the number of susceptibles. By contrast the Flux model shows non-monotonic behavior due to back-filling of susceptibles from the smaller outbreak in location 2 traveling back to location 1. **A**: Short travel duration regime ($\phi_i = 1, \tau_{i,j} = 10$); low transmission in location 2 ($\beta_2 = 0.5$) **B**: Short travel duration regime ($\phi_i = 1, \tau_{i,j} = 10$); high transmission in location 2 ($\beta_2 = 2.5$) **C**: Long travel duration regime ($\phi_i = 0.01, \tau_{i,j} = 0.1$); low transmission in location 2 ($\beta_2 = 0.5$) **D**: Long travel duration regime ($\phi_i = 0.01, \tau_{i,j} = 0.1$); high transmission in location 2 ($\beta_2 = 2.5$)

We therefore model the rates at which people travel from $i$ to $j$ by decomposing each rate into two components. First, a person decides to leaves home at a rate $\pi_i$. Second, the traveler decides on their destination region $d$ with probability $(\mathbf{P}(i \to d \mid i))$. Lastly, each traveler chooses a destination pixel $j$ within the chosen destination region $d$ $(\mathbf{P}(d \to j \mid i, d))$. Combining all three components together:

$$\phi_{i,j} = \pi_i \mathbf{P}(i \to d \mid i)\mathbf{P}(d \to j \mid i, d) \qquad [1]$$

We model the rate at which people leave home as $\pi_i = \mathbf{P}(\text{Travel} \mid i)/60$, the probability that an individual living at $i$ traveled away from home during the 60 days covered by the MIS study period, divided by the length of the study period. For each pixel, the probability of leaving $(\mathbf{P}(\text{Travel} \mid i))$ was estimated using the same geostatistical techniques used to estimate prevalence, first reported in (6) and reproduced with permission in Figure 4 B.

We model the conditional probability distribution $\mathbf{P}(i \to d \mid i)$ as a multinomial probability distribution. The MIS data include counts of the number of times a traveler from $i$ traveled to one of the seven destination regions. From the counts data, we fit a multinomial regression using the `nnet` package in `R`. The regression model included as additional covariates the region on Bioko Island where $i$ is located; the population $N_i$; and the distance between $i$ and the centroid of each destination region. (Other covariate combinations were tested and found to negatively effect the model's performance, as assessed using AIC.)

Data on travel to specific destination pixels within each destination region was not available from the MIS. For this reason, we make the assumption that a traveler is equally likely to travel to visit any single individual living in any destination region. From this assumption, the probability of traveling to each pixel is proportional to the population of $j$ within $d$: $\mathbf{P}(\mathbf{d} \to \mathbf{j} \mid \mathbf{i}, \mathbf{d}) = N_j / \sum_{k \in d} N_k$. Varying this assumption, for example to allow travelers to visit each destination pixel equally, makes no noticeable change to the results reported here.

**B. Travel Duration.** In addition to questions about travel duration, in 2018 MIS respondents reported on how much time they spent away from home if they had traveled. We first split the data into travel on-island and travel off-island, noting that off-island trips are often longer and require traveling by boat or airplane. We used maximum likelihood methods to fit an exponential decay model to each subset of the trip duration data, where the exponential decay model reflects the mechanistic Simple Trip model's assumption of a constant rate of returning home. Average duration of on-island travel was estimated to be $\tau_{i,\text{EG}}^{-1} = 10.3$ days. Average duration of off-island travel was estimated to be $\tau_{i,\text{BI}}^{-1} = 21.2$ days.

### 3. Variables and Notation

**Table 1. Movement Model Parameter Definition**

| Parameter | Interpretation | Model |
|---|---|---|
| $f_{i,j}$ | Rate at which member of $i$ travels $i \to j$ | Flux model |
| $\phi_{i,j}$ | Rate at which a resident of $i$ travels $i \to j$ | Simple Trip model |
| $\tau_{i,j}$ | Rate at which a visitor to $j$ returns home to $i$ | Simple Trip model |
| $\psi_{i,j}$ | Average fraction of time spent in location $j$ for residents of location $i$ | Simple Trip model |

**Table 2. SIR and SIS Model State Variables and Parameters**

| State Variable or Parameter | Interpretation | Value (if constant) |
|---|---|---|
| $S$ | Number of susceptible hosts | |
| $I$ | Number of infected/infectious hosts | |
| $R$ | Number of removed/recovered hosts | |
| $N$ | Total number of hosts in the metapopulation | |
| $\beta$ | Transmission intensity, per-contact rate of infection | |
| $\gamma$ | Recovery rate | $\gamma = 1$ |

**Table 3. Ross-Macdonald Model State Variables and Parameters**

| State Variable or Parameter | Interpretation | Value (if constant) |
|---|---|---|
| $X$ | Number of infected/infectious human hosts | |
| $N$ | Total number of humans | Assumed to be constant |
| $r$ | Rate of recovery | $r = 1/200$ day$^{-1}$ |
| $a$ | Mosquito bites per human per day | $a = 0.27$ |
| $b$ | Mosquito-to-human transmission efficiency | $b = 0.55$ |
| $Z$ | Number of infectious mosquitoes | |
| $M$ | Total number of mosquitoes | Assumed to be constant in time |
| $c$ | Human-to-mosquito transmission efficiency | $c = 0.15$ |
| $g$ | Rate of mosquito death | $g = 0.1$ |
| $e^{-gn}$ | Fraction of mosquitoes who survive the extrinsic incubation period (EIP) | $n = 11$ days, $e^{-gn} \approx 0.33$ |

### 4. Guide to Supplementary Code and Data

Code and data supporting the analysis performed in the Malaria Modeling Example section of the main text may be found at https://doi.org/10.6084/m9.figshare.12084831.v2

The data files included in the linked repository are as follows:

1. `Dataset_S1_raw_survey_data.csv` - Travel survey data, counting number of trips taken by residents of each area on Bioko Island. Derived from the Malaria Indicator Survey Data (2–4).
2. `Dataset_S2_modeled_PR_and_travel.csv` - For each area on Bioko Island, shows estimated prevalence (PR) (6) and estimated probability of traveling to each destination region
3. `Dataset_S3_travel_duration_data.csv` - Records the duration (in nights) of trips reported in the survey data (2–4) and whether those trips were to off-island (mainland Equatorial Guinea) or on-island (elsewhere on Bioko Island)
4. `Dataset_S4_time_at_risk_matrix.csv` - The time at risk matrix, for the purpose of calculating the values of $R_0$ for the Simple Trip model.

Additionally, we have attached two R Markdown code notebooks which may be used along with the datasets in the repository to reproduce the analysis described in the Malaria Modeling Example section of the main text and Supplementary Information section 2.

1. `Travel_Destination_Choice_Model.Rmd` - used to model the probability distribution of traveling to each possible destination from each possible origin. Takes `Dataset_S1_raw_survey_data.csv` as input and generates the travel destination probability data found in `Dataset_S2_modeled_PR_and_travel.csv`.
2. `Comparing_Metapopulation_Dynamics_companion_code.Rmd` - Takes `Dataset_S2` and `Dataset_S3` as input. Used to model travel duration, the time at risk matrix (`Dataset_S4_time_at_risk_matrix.csv`), and $R_0$ for both the Flux and Simple Trip models


1. SY Kang, et al., Spatio-temporal mapping of Madagascar's Malaria Indicator Survey results to assess Plasmodium falciparum endemicity trends between 2011 and 2016. *BMC Medicine* **16**, 1–15 (2018).
2. The Government of Equatorial Guinea, Medical Care Development International (MCDI), The bioko island malaria control project malaria indicator survey (MIS) 2015, (Medical Care Development International), Technical report (2015).
3. The Government of Equatorial Guinea, Medical Care Development International (MCDI), Bioko island malaria control project iii - malaria indicator survey (MIS) 2016, (Medical Care Development International), Technical report (2016).
4. The Government of Equatorial Guinea, Medical Care Development International (MCDI), The bioko island malaria control project malaria indicator survey (MIS) 2017, (Medical Care Development International), Technical report (2017).
5. PW Gething, et al., A new world malaria map: Plasmodium falciparum endemicity in 2010. *Malar. J.* **10**, 378 (2011).
6. CA Guerra, et al., Human mobility patterns and malaria importation on Bioko Island. **10** (2019).
